# Pre-analytical delay of blood cultures: An observational study with an appraisal of the effects of laboratory centralisation

**DOI:** 10.64898/2026.03.19.26348778

**Authors:** Malila R Noone

**Affiliations:** Independent researcher

## Abstract

Aims The United Kingdom Standards for Microbiology Investigations (UK SMI) recommends incubating inoculated blood cultures in an automated analyser as soon as possible and within 4 hours of collection. This observational study records compliance with a four hour standard by microbiology services across NHS England. The impact of laboratory centralisation on compliance rates was appraised.

**Methods:** Freedom of Information requests were submitted to 116 National Health Service Trusts/administrative units in England requesting retrospective audit data showing compliance with the standard over a single year. Additional information requested relating to service configuration and laboratory costs was supplemented by data available in the public domain.

**Results:** Responses were received from 89 Trusts (76.7%) comprising 149 acute hospitals. Compliance was poor with only four hospitals (2.7%) showing full compliance with the recommended standard. There was wide variation in compliance rates between hospitals across the region and hospitals within the same Trust and within Trust networks. Laboratory centralisation was not accompanied by a cohesive strategy which addressed the particular needs of a microbiology service. These changes hinder the optimal management of blood cultures.

**Conclusions:** Pre-analytical delay of blood cultures impedes timely reports which are critical to the effective management of sepsis while limiting the emergence of antimicrobial resistance. The need for a more comprehensive microbiology service in each acute care setting can be rationalised in view of the continued high prevalence of sepsis and the increasing emergence of antimicrobial resistance.

## INTRODUCTION

Sepsis is a complication of infection which is associated with a high mortality rate and life changing after-effects. These have been brought to public attention by Surviving Sepsis and other campaigns [1–3] and rising rates of sepsis are reported [4, 5]. While the clinical diagnosis of sepsis is based on international consensus guidelines [6], blood culture remains the gold standard test which confirms the infection and identifies the antimicrobial agents most likely to be effective [7]. Early recognition of sepsis and prompt initiation of antimicrobial therapy is crucial to survival, and the current practice of administering a cocktail of antimicrobial agents on an empirical basis, has proved to be effective [8–10]. Early pathogen identification is critical as it may prompt a lifesaving switch to more effective agents if initial empirical therapy is deemed to be sub-optimal. The likelihood of this occurrence is increasing, with the growing prevalence of antimicrobial resistance (AMR) [11, 12]. Although effective, the use of broad spectrum antimicrobial therapy has itself been linked to the emergence of AMR [13] and current antimicrobial stewardship programs strongly support early de-escalation to the most effective narrow spectrum agents [14]. Current continuous-monitoring automated blood culture systems show increased sensitivity and can provide provisional reports within 08-18 hours of procuring a blood culture if collection and management of the specimen is optimised [15]. Poor specimen management will hinder de-escalation, prolong broad spectrum antibiotic therapy and foster the emergence of AMR and other iatrogenic harms [16, 17]. Early identification of the pathogen has added clinical benefits. It may point to the probable source of infection and, since mortality risk and clinical progress varies across organisms, also serves as a prognostic indicator guiding the use of adjunctive therapeutic agents and clinical interventions [18]. When reports of pathogens and their susceptibilities are expedited, there is a demonstrable impact on mortality rates and health care costs [19, 20]. A negative result is also of considerable value as it promotes withdrawal of antimicrobials altogether. However, unless the validity of a negative report is assured by documentation of optimal specimen management, a negative result may be discounted, leading to needlessly prolonged antimicrobial therapy and its associated harms.

In summary, timely blood culture results support the effective management of sepsis and limit the emergence of AMR.

Pathogen survival in blood culture media is variable and affected by many factors such as species characteristics and holding temperatures. The effect of delay on survival is however recognised as an important controllable factor and the United Kingdom Standards for Microbiology Investigations (UK SMI) recommends incubating inoculated blood cultures in an automated analyser as soon as possible and within 4 hours of collection [21]. Minimising pre-analytical delay (PAD) provides early results, but also increases detection rates and decreases time to detection by the analyser. Conversely, pre-analytical delay, as occurs outside laboratory operating hours, results in a reduced yield [22–25]. It has even been proposed that the delay in handling blood cultures taken over the weekend when laboratory services were reduced, may be linked to the increased mortality observed over the weekend and branded the ‘’weekend effect’’ [26, 27]. These reports highlight the impact of microbiology service schedules on blood culture results.

Microbiology services have been subject to major changes in England. In 1990, acute care hospitals in England were set up as independent and competitive administrative units termed Trusts. Each Trust provided acute care laboratory services for patients in hospital as well as a primary care community service for General Practitioners (GPs) in a designated geographical area. When Trusts merged, hospital management was integrated but a laboratory was initially retained by each hospital. In 2006, an Independent Review found England’s NHS pathology service to be one of the best value systems in the world with expenditure per capita on in-vitro diagnostics about half that of equivalent countries in Europe, and a quarter of that in the USA [28]. Nevertheless, in 2008, “back office and pathology services” (categorised as being comparable) were ear-marked for speedy centralisation [29]. Centralisation was driven by the potential for savings, with a target reduction of global pathology expenditure (expenses for all pathology specialities) from approximately 4% of total Trust expenditure to <1.6%. The ‘hub and spoke’ model recommended was expected to achieve benefits through economies of scale by concentrating ‘non-urgent’ work at a centralised ‘hub’. Microbiology specimens were deemed non-urgent and ‘spoke’ hospitals transferred them to the hub while retaining their clinical (usually medically qualified) microbiologists on site. In spoke hospitals, ‘essential services laboratories’ (ESLs) provided only those tests which were defined as time critical and requiring an analytical turn–around-time (TAT) of four hours. Although analytical TATs of microbiological specimens range from three to five days, they are also time critical and international guidelines recommend they are processed “as soon as possible” [30]. The diversity of infectious agents and specimen types preclude precise TATs for microbiological specimens and leaves the recommendation ‘’as soon as possible’’ open to interpretation or disregarded altogether. This was a cause for concern and was considered ‘a step too far’ when centralisation of microbiology services was introduced in England [31].

This retrospective observational study documents compliance with the PAD standard by microbiology services in England during the financial year 2022/23. The changes accompanying centralisation and their impact on microbiology services are appraised. Wider clinical implications and governance issues are also examined and future direction considered.

## MATERIALS AND METHODS

### Study Design

Freedom of Information requests [Appendix 1] were submitted to acute-care non-specialist Trusts which were listed in the NHS England provider directory. The submission was addressed to the designated FOI officer in each of 116 Trusts, with the expectation of collaboration with the Trust’s clinical microbiologist. The principal enquiry related to the percentage of blood culture sets incubated within four hours of specimen procurement at each acute hospital within the Trust in the financial year 2022/23. The reported information relating to service configuration was supplemented by data available in the public domain. The standard opening hours of each laboratory was not requested, as compliance with the PAD standard only relates to incubation facilities rather than specimen processing facilities. Global pathology cost as a percentage of total Trust costs for the same year was also requested. This precise cost of the laboratory service is not usually displayed in the Trust annual financial reports which are available to the public.

The UK Freedom of Information Act 2000 provides right of access to information held by public authorities including the NHS, and Trusts are legally obliged to respond to requests. A public body may refuse to release information which is deemed ‘sensitive’, is not readily available or can be found elsewhere, but any information released in response to a FOI request is deemed to be factually correct. The information, once released, is in the public domain and may be shared with appropriate safeguards. The data reported here do not contravene copyright restrictions and personal data protection guidance and preclude identification of individual Trusts. Further data cannot be provided as it may lead to identification of individual Trusts.

## RESULTS

Of the 116 Trusts approached, 15 did not reply. They were not pursued due to awareness that administrative changes and clinical service emergencies may prevent prompt responses to FOI requests. A further 12 Trusts stated that they did not have the data requested and suggested referral of the request to their off-site service provider. Since an off-site provider would not be responsible for the pre-analytical management of specimens, an appeal was made for the Trust clinical microbiologist to be made aware of the FOI request. This did not change the stand taken.

### Service configurations

Data are presented for 89 Trusts. The 89 Trusts managed 146 acute hospitals providing 24 hour acute-care services and submitting more than 500 blood cultures annually. Service configurations varied widely. (Table 1)

**Table 1:**
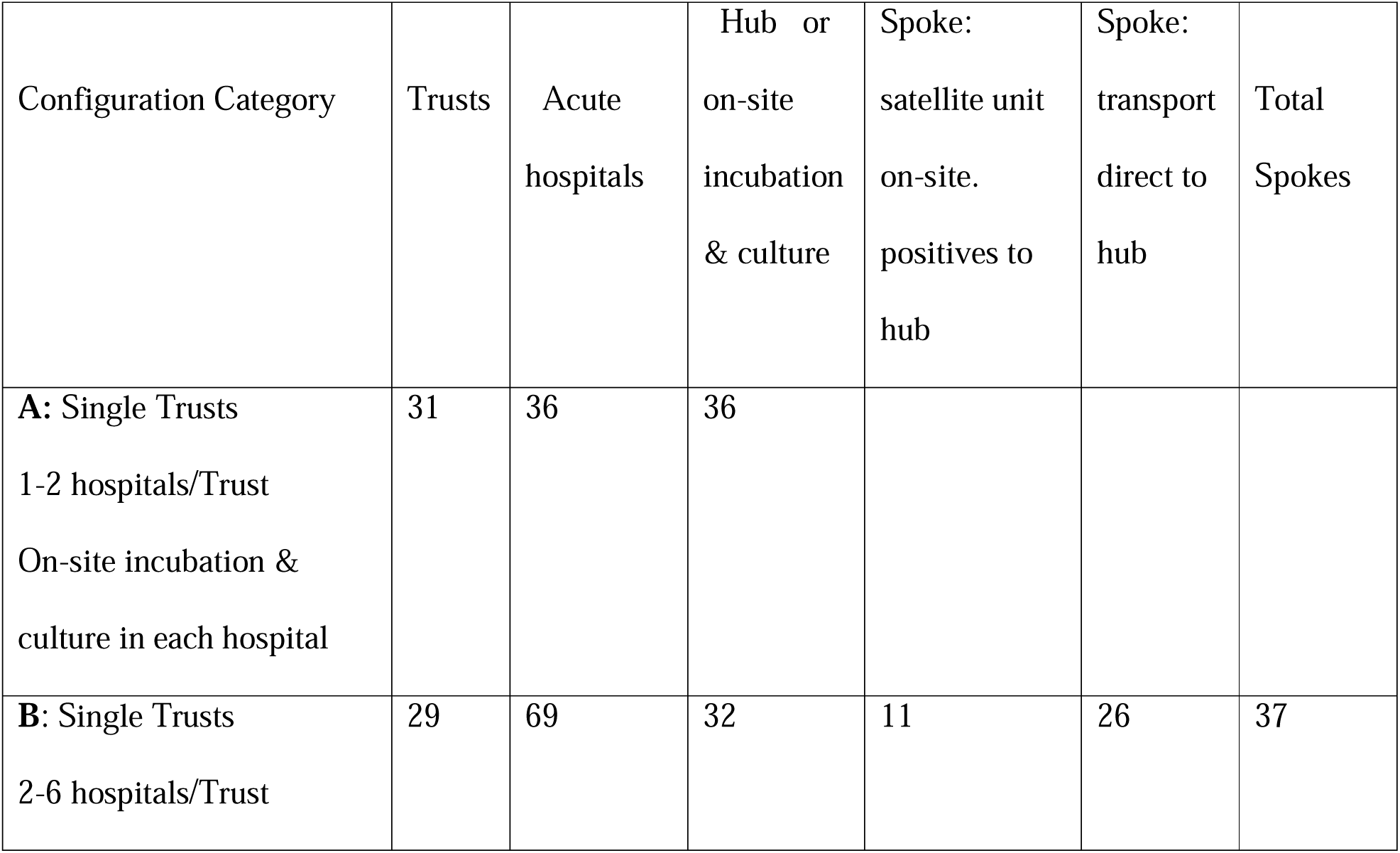
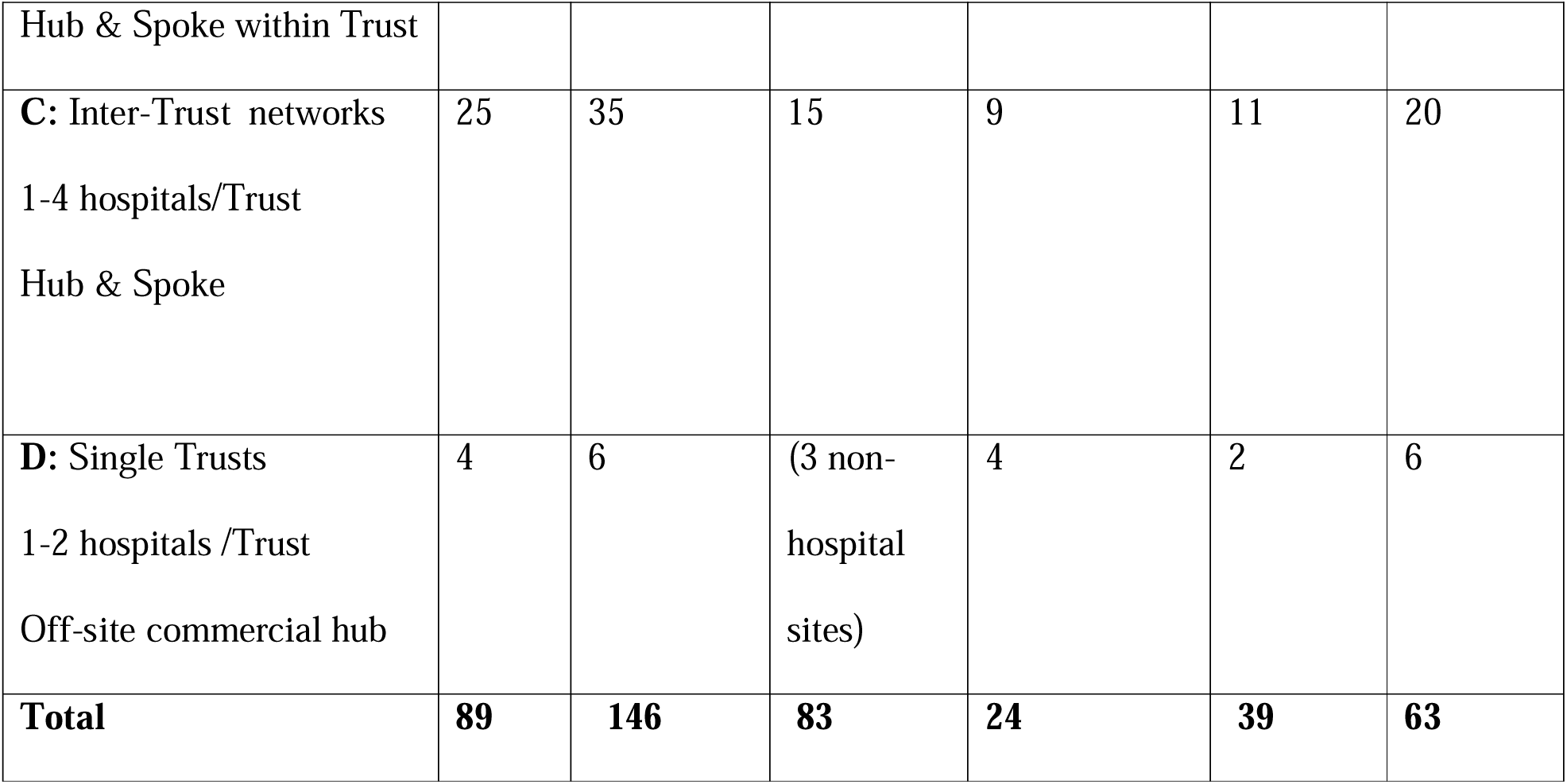
Service configurations in 89 Trusts.

Thirty-one Trusts managing 1-2 hospitals each had retained arrangements for incubation and processing of blood cultures on-site in each hospital. Fifty-eight Trusts (65%) had established hub and spoke arrangements either within their own Trust, as inter-Trust networks, or by using commercial off site providers. There were 63 spoke hospitals. Of these, 39 transported specimens directly to the hub while 24 had satellite units for on-site incubation of cultures. Only bottles flagging positive at the latter sites were transported to the hub laboratory to be processed. In one of these spoke hospitals, specimens flagging positive within 18 hours were tested using a BioFire Blood Culture Identification Panel before transfer to the hub. Spoke to hub distances for the 63 spoke hospitals ranged from 1.9-40miles (median 10miles).

### Compliance with the Pre-Analytical Delay standard

Only 53 of 89 Trusts (59.5%) had compliance data measured on at least one occasion. Only 14 Trusts (15.7%) reported undertaking regular audit. As there were disparities in compliance rates between hospitals in a single Trust and between hospitals within a single inter-trust network, compliance rates are presented for the 146 hospitals. (Table 2)

**Table 2:** Compliance with pre-analytical standards in 146 hospitals.

|  | 100% | 80-99% | 50-79% | <50% | No data | Total |
| --- | --- | --- | --- | --- | --- | --- |
| Hubs or on site culture | 3 | 24 | 25 | 9 | 22 | 83 |
| Spokes with satellite unit | 1 | 12 | 3 | 4 | 4 | 24 |
| Spokes –transport only |  | 6 | 4 | 19 | 10 | 39 |
| **Total** | **4** | **42** | **32** | **32** | **36** | **146** |

Full compliance was reported by only four of hospitals (2.7%). A further 42 hospitals (28.7%) reported that >80% of specimens were incubated within four hours (>80% compliance). This rate of compliance is suggestive of incubation of cultures beyond routine laboratory hours or the use of satellite units, as it was reported that when incubation was restricted to routine laboratory hours, only approximately 50% of specimens achieved incubation within the four hour standard.

Thirty-six Trusts (40.4%) had no compliance data for 2022/23. Reasons given for the lack of data included inability to access or interrogate the computer system and inability to change computer settings. This included the inability to record the time of specimen procurement although this is a legal requirement and recorded on both the request form and specimen label. The interval between time of specimen procurement and time of incubation was not routinely included in reports. Several Trusts reacted when delays were noticed, but only one Trust reported inclusion of an explicit comment in the final report indicating that the blood culture was received in the laboratory more than four hours after it was taken and that the delay could reduce the chance of growing relevant pathogens.

### Cost of the laboratory Service as a percentage of Trust cost: Table 3

Costs were provided by 71 Trusts (79.7%). Eighteen Trusts withheld cost data citing it commercially sensitive or confidential. (Table 3)

**Table 3:** Costs by Configuration: Interquartile range and median.

| Configuration Category | Trusts | Cost reports | Costs withheld | IQ Range/ median |
| --- | --- | --- | --- | --- |
| A: On- site incubation and culture | 31 | 28 | 3 | 2.9-4.0/ 3.19 |
| B: Hub & Spoke within Trust | 29 | 23 | 6 | 3.0-4.6/3.3 |
| C:Hub & Spoke Inter-Trust | 25 | 17 | 8 | 3.0-4.7/3.7 |
| D:Off-site commercial | 4 | 3 | 1 | N/A |
| **Total** | **89** | **71** | **18** | **2.95-4.31/3.3** |

Overall, the reported interquartile range of costs was 2.95-4.31 (Median 3.3%). Hub and spoke arrangements did not appear to confer a cost advantage over Trusts which had retained on-site incubation and culture (Median 3.19).

## DISCUSSION

The vital importance of the early recognition of sepsis and the prompt initiation of antimicrobial therapy has been widely publicised as being crucial to survival. The critical role played by clinical microbiology services in supporting the management of sepsis and reducing the emergence of antimicrobial resistance is less well recognised and has not been monitored. Compliance with the recommended standard which minimises the pre-analytical delay of blood cultures is an auditable quality measure of a microbiology service and reflects its ability to support the effective management of sepsis. The study shows that only four of 146 hospitals (2.7%) in 89 Trusts in England showed full compliance with this key quality indicator. A further 42 hospitals (28.7%) reported that >80% of specimens were incubated within four hours (>80% compliance). It may be argued that this rate of compliance is acceptable, but it is less than ideal and supports service improvement.

This report is unique in that it relates to microbiology services across a single region served by a single healthcare provider (NHS England) which encouraged laboratory centralisation. The impact of laboratory centralisation on compliance rates and on wider healthcare and governance issues can be judged using the incidental data obtained. The factors highlighted below relate in particular to planning prior to the implementation of centralisation, and the consideration given to the particular requirements of the microbiology service.

### Service configurations

Although some variation in service configuration is expected, in keeping with geographical contours and the distribution of urban concentrations, the extremes observed here points to the lack of a cohesive strategy when centralisation was imposed. The disparities in quality standards between hospitals in the region, and often within a single management entity, further indicate that decisions relating to the service and quality targets were left to individual Trusts. A core strategy with an obligatory quality framework might have facilitated prospective review of the effects of centralisation but this has not been undertaken. A proactive region- wide review of post-centralisation microbiology services has been undertaken elsewhere [32, 33].

### Hub and spoke systems

This recommended system applied to 58 Trusts (65%). Sixty three spoke hospitals had suffered the loss of their on-site microbiology laboratories and their specialist Biomedical Scientists (BMS). Hypothetically, some of the financial savings thus released might have been available for investment in robust transport facilities and enhanced out of hours (OOH) services. There does not however appear to have been a mandate for this. Specimen transport arrangements appear less than adequate as, of 39 spoke hospitals relying on transport, only 6 (15.3%) showed >80% compliance. Limited enhancement of OOH services was seen in 42 hospitals (28.7%) which showed >80% compliance. Compliance of 80% is easily achieved by a single negotiated attendance by a BMS between 10pm and midnights as only 9% of attendances at NHS emergency centres occur between the hours of midnight and 7am, with a fall in blood culture numbers during these hours (personal experience).

### Information Technology & Audit

Regular audit of procedural standards is recognised as good laboratory practice by the Royal College of Pathologists (RCPath) [34]. Although laboratory centralisation was accompanied by investment in information technology (IT) infrastructure, auditable standards for microbiology do not appear to have been incorporated into laboratory systems. Thirty-six Trusts (40.4%) had no compliance data for 2022/23 and only 14 Trusts (15.9%) reported an ability to undertake regular audits. The inherent complexity and changing needs of microbiology present a challenge and resolution is more problematic when IT support is off-site or outsourced.

### Savings

Centralisation was driven by the demand for cost savings but the target reduction of pathology expenditure to <1.6% of Trust expenditure has not been realised. Although further examination will be required to analyse the distribution of costs by department, the potential contribution of the microbiology laboratory to the savings agenda is likely to be limited. Centralising serological specimens which benefit from automation promise savings but these may not be realised due to costs arising from transport, bio-hazard regulations and tracking. Economies of scale do not readily translate to specimens needing labour intensive manual procedures and transport delays affect the quality of these specimens. The NHS should be able to take advantage of discounts associated with bulk purchasing and leasing agreements across Trusts and, except in the case of specialised investigations, should be able to retain microbiology tests on site. It is noteworthy that hub and spoke arrangements did not appear to confer a cost advantage over Trusts which had retained microbiology services on-site.

### Governance & quality management

Responsibility for maintaining and auditing quality clearly lies with the designated hospital clinical microbiologist and this has ethical implications [35]. Referral of the FOI request to the off- site service provider on 12 occasions hints at a lack of direct control, a lack of ownership, disengagement with quality issues or a sense of alienation following the move of the hospital laboratory service off-site. The lack of mandatory status for UK SMIs may also have disempowered clinical microbiologists and curtailed their ability to secure the investment and support required to upgrade their service. Although Laboratory centralisation has been undertaken in many countries in pursuit of financial savings, there are few reports of its impact on subsequent laboratory performance and quality [33].

### Wider aspects of healthcare

There have been no proactive measures in place to monitor the impact of laboratory centralisation on wider aspects of healthcare such as duration of stay, antibiotic use, healthcare associated infections and the emergence of antimicrobial resistance. Although healthcare outcomes are reported by Trusts, they are rarely delineated by hospital and there is no reference to the local configuration of microbiology services [36]. Consequently, iatrogenic patient harms and in particular, the emergence of AMR may not be recognised as attributable to the laboratory shortcomings which may have followed centralisation in England. Elsewhere, the impact of microbiology laboratory centralisation on antimicrobial stewardship and hospital acquired infections and their associated costs have been reported [37–39].

### Limitations of recent modifications

An NHS England directive to all Trusts encouraged compliance with the PAD standard in the interests of improving antimicrobial stewardship and patient outcomes from sepsis [40] and a limited reversal of centralisation has been undertaken by the installation of satellite units in hospitals which had lost on-site microbiology facilities. Prompt incubation of blood cultures is, however, only the first step in the pathway to providing speedy results. If positive bottles are not processed swiftly, delays in time to Gram stain and time to antimicrobial test results will continue to cause patient harm [41]. An adequate response to the Surviving Sepsis campaigns requires a seamless and speedy pathway from specimen collection to the final report. The use of molecular tests on positive bottles and on EDTA blood samples will speed up the management of sepsis but only in relation to most common pathogens. These new diagnostic systems do not support centralisation of microbiology facilities as they require effective on-site clinical liaison which is more difficult when the service is centralised [42]. Furthermore, new technologies cannot entirely replace standard diagnostic microbiology which is critical for the detection of rare and emerging pathogens and unusual antimicrobial resistance patterns. In addition to blood cultures, specimens from associated sites also warrant prompt attention as they may yield the only positive isolate from a patient with sepsis.

### Conclusions and future direction

Appraisal of the changes which accompanied laboratory centralisation in England indicates that the particular needs of a microbiology service were overlooked. These changes hinder the optimal management of blood cultures which in turn impacts on wider healthcare issues. The future direction of microbiology services in England should be considered in the context of the continued high prevalence of sepsis and the emergence of antimicrobial resistance. Where laboratory centralisation has been implemented in other countries, there has been concern about its effect on microbiology services, and the need for a more comprehensive service in each acute care setting has been rationalised [37–39]. Wider clinical needs, control of infection requirements, antibiotic stewardship, and the ability to respond to national demands such as outbreaks of infectious disease support this view and indicates that microbiology should be recognised as an essential 24-hour on-site service. Mandatory institution of National good practice recommendations and associated standards [43] are likely to be cost effective if the impact on patient survival and indirect savings in health care costs are taken into account. There will also be considerable community benefit in terms of containing the emergence of antimicrobial resistance.

## Data Availability

All relevant data are contained within the manuscript. Data reported here do not contravene copyright restrictions and personal data protection guidance and preclude identification of individual Trusts. Further data cannot be provided as it may lead to identification of individual Trusts.

## Acknowledgements

The microbiologists who responded to the questionnaire especially those who included informative comments.

Application for ethical approval was not considered necessary as patient data were not requested. The author received no funding for this study and has no competing interests.

This study is unique in that it relates to microbiology services across a single region served by a single healthcare provider (NHS England). There have been no previous reports relating to this subject.

## Appendix 1: FOI blood culture request

This is a FOI request relating to the microbiology laboratory service and blood cultures.

The data should be easily available by interrogating laboratory information systems or from audit reports. I should be grateful for a response in the form detailed below.

Please let me know if any of the questions are ambiguous or unclear.

The data will not be used in a way which will identify an individual Trust. Thank you for your time.

## FREEDOM OF INFORMATION REQUEST: BLOOD CULTURES

1. Please table the hospitals or other acute sites currently submitting blood cultures and for each site-

i. Are blood cultures incubated and/or processed on site or sent to a microbiology laboratory off site?
ii. If incubated or processed off site, what is the distance in miles between the site and the offsite laboratory?
iii. How many blood culture sets were submitted from each site in 2022/23?
iv. Number of bed days for all patients per site

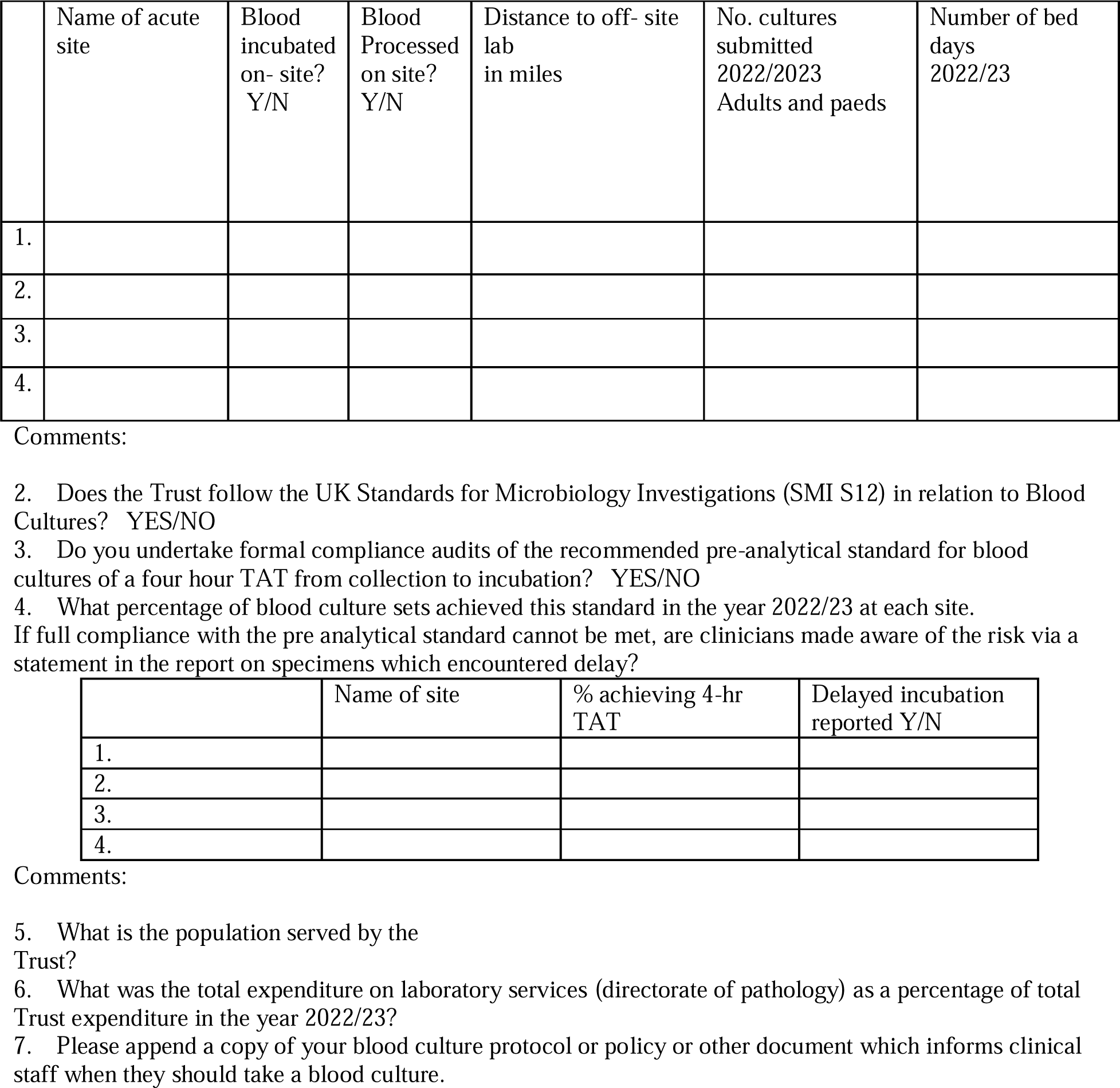

## Notes

### Competing Interest Statement

The authors have declared no competing interest.

### Funding Statement

The author(s) received no specific funding for this work.

### Summary of Updates

This version presents the same data and results as before. The views and opinions presented in relation to the significance of pre-analytical delay of blood cultures and the impact of laboratory centralisation on microbiology services is better supported, with appropriate reference to the current literature. This version manuscript is more concise.

